# Feasibility of Distributed Wearable Ultrasound for Detecting Neuromuscular Coordination Differences After Knee Injury

**DOI:** 10.64898/2026.10.18.26365835

**Authors:** Erica L. King, Morgan A. Lamarre, Ahmed Bashatah, Abhishek Aher, Gabriel Gibson, Tina Hajialiyan, Faith S.A. Brown, Qi Wei, Siddhartha Sikdar, Margaret T. Jones, Parag V. Chitnis

## Abstract

**Objective:** The aim of the current study was to evaluate the feasibility of a multi-channel distributed wearable ultrasound (D-WUS) system capable of simultaneously probing dynamic muscle activity across multiple anatomical sites to assess neuromuscular coordination during physical activity.

**Methods:** Twenty NCAA Division I women athletes (8 with prior knee injury) performed countermovement jumps (CMJs) on dual force plates while wearing a four-channel D-WUS system targeting the vastus lateralis (VL) and vastus medialis oblique (VMO) in both limbs. Real-time M-mode ultrasound data were processed using a composite factorization approach integrating nonnegative matrix factorization and principal component analysis (CF-PCA) to extract dominant muscle-contraction patterns. Correlation and timing metrics between VL and VMO were compared across injury status.

**Results:** CMJ performance metrics (jump height, RSImod) did not differ between groups (p > 0.05). However, D-WUS revealed significant differences in muscle coordination in the right limb: VL–VMO correlation (p = 0.029) and timing symmetry (p = 0.019) were reduced in participants with prior knee injury; left-limb differences were not significant. Right-limb timing symmetry was negatively correlated with SMFA score (ρ = −0.48, p = 0.033). Functional survey scores (SMFA) also differed significantly (p = 0.001), while VISA-P scores did not.

**Conclusion:** These preliminary findings provide feasibility-level evidence that distributed wearable ultrasound can detect limb-specific neuromuscular coordination differences not reflected in force-plate biomechanics, motivating larger prospective studies to establish reliability and clinical utility.

**Significance:** With further validation, this approach may offer a scalable, objective means of monitoring muscle contraction during rehabilitation.

## Introduction

Once a musculoskeletal injury (MSKI) occurs, neuromuscular function rapidly degrades [1]. Rehabilitation following MSKIs centers on restoring normal muscle activation patterns and neuromuscular coordination to ensure a safe and effective return to activity [2], [3]. Proper muscle function is essential for coordinated movement, and deviations from normal activation patterns can contribute to compensatory movement that may increase the risk of re-injury or hinder optimal performance [4], [5], [6].

Understanding and monitoring synergist muscle activation patterns is crucial for developing effective rehabilitation protocols, particularly in restoring neuromuscular coordination after knee injury [1]. Neuromuscular coordination plays a vital role in post-injury rehabilitation, as deficits in muscle activation timing and synergy can lead to altered movement mechanics and suboptimal performance [3], [4], [8], [9], [10].

For instance, when subjects recovering from knee injuries are undergoing rehabilitation, they often exhibit altered jump mechanics, asymmetrical force distribution, and delayed muscle activation patterns, clear indicators of neuromuscular deficits [10], [11]. Therefore, the countermovement jump (CMJ) is commonly used in rehabilitation protocols to assess lower-limb force production, movement symmetry, and neuromuscular coordination [12], [13], [14].

While bilateral force plates offer whole-body neuromuscular assessment, they do not capture muscle-level activation patterns, which is a critical limitation when subtle compensatory actions or asymmetries persist post-injury. Muscle-level symmetry assessments are essential to detect imbalances that may contribute to dysfunction or increased re-injury risk [15], [16]. Current methods (e.g., visual assessment, palpation, manual muscle testing) are inherently subjective and often fail to detect nuanced neuromuscular deficits [17].

Surface electromyography (sEMG) has traditionally been used to assess muscle activity at the tissue level, but it suffers from limitations such as signal noise, muscle crosstalk, and poor sensitivity to deep muscle groups, especially during dynamic movements [18], [19]. Similarly, conventional handheld ultrasound systems provide valuable anatomical and functional insights but are constrained by operator dependence, single-site imaging, and impracticality for continuous monitoring during performance tasks [20], [21], [22], [23], [24]. To comprehensively capture muscle synergies, including the interplay between multiple muscles, there is a growing need for a multi-site wearable ultrasound system that can simultaneously track muscle activation patterns in different muscles, providing a more holistic assessment of neuromuscular coordination during dynamic movements.

Current applications of wearable ultrasound have primarily focused on cardiovascular and pulmonary function, as well as exoskeleton and prosthesis control at a single site or several sites in an unsynchronized manner [25], [26], [27], [28], [29], [30], [31], [32]. Nevertheless, wearable ultrasound holds great promise for simultaneously imaging different muscle groups contributing to force production during movement, thereby enabling a comprehensive and systematic assessment of muscle function [26], [27]. Achieving this goal requires a miniaturized system capable of distributed imaging across multiple muscle groups concurrently.

Motion-mode (M-mode) ultrasound imaging offers a viable alternative to conventional brightness-mode (B-mode) imaging for measuring real-time muscle-contraction. However, traditional ultrasound systems extract M-mode signals by acquiring standard B-mode image sequences and sampling a single image line from each frame. Furthermore, extracting meaningful functional measures from M-mode images remains challenging due to fast-moving complex tissue interfaces during highly dynamic physical activity [33].

To overcome these challenges, we integrate two strategies: a custom M-mode ultrasound device based on time-delay spectroscopy built from the ground up for directly providing multi-site (multi-channel) M-mode images in a compact wearable form-factor [26], [29] amenable to wireless implementation; and a composite signal processing pipeline that combines Nonnegative Matrix Factorization (NNMF) and Principal Component Analysis (PCA) for extracting muscle-contraction signals from dynamic M-mode images. NNMF identifies dominant, nonnegative activation patterns, while PCA reduces dimensionality to isolate key components explaining signal variance. Similar approaches have been used in previous research with NNMF-only for accurately extracting gait phases from clinical 2-dimensional ultrasound images [34]. PCA has been used in ultrasound research to classify hand gestures in signals obtained by A-line ultrasound scans [35].

However, these signal processing schemes have inherent limitations. Traditional NNMF approaches are limited in that they do not account for statistical dependencies among extracted components or reveal underlying structural relationships within the data. The results of NNMF are also influenced by input parameters including the number of factors selected for the decomposition. Furthermore, the resulting parts-based representation can sometimes appear global rather than localized, depending on the characteristics and complexity of the dataset [36], [37].

PCA also has limitations in that it assumes that the directions of maximum variance in the data correspond to meaningful structure, which may not hold true for complex or nonlinear relationships [37]. Additionally, PCA is sensitive to scaling and noise, and it does not capture temporal or spatial dependencies within the data, limiting its ability to model intricate patterns [37].

In this study, these methods are combined in a composite factorization approach, Composite Factorization PCA (CF-PCA), in which NNMF is run across a range of factorization ranks and PCA is applied to the concatenated output. This reduces sensitivity to the choice of rank and to random initialization and does not require labeled training data. CF-PCA has been benchmarked against expert-annotated ground truth using this wearable hardware [57].

The vastus lateralis (VL) and vastus medialis oblique (VMO) are the primary quadriceps muscles governing patellar tracking during knee extension. Imbalanced timing between VL and VMO activation is a well-documented feature of patellofemoral pain syndrome and has been reported following knee injury [16], [50], [51]. The CMJ imposes high eccentric and concentric demands on the extensor mechanism, making it a suitable task for detecting coordination deficits between these muscles. Both muscles are superficial and accessible to surface-mounted ultrasound transducers, making them practical targets for the distributed wearable system described here.

In this feasibility study, we hypothesized that this system could detect subtle deficits in muscle activation timing and symmetry in individuals with a history of knee injury, which are not captured by conventional biomechanical tools such as force plates. By pairing wearable ultrasound with established rehabilitation and assessment protocols, this work takes an initial step toward a scalable, objective method for analysis of neuromuscular function.

## Methods

Participants were recruited to perform maximum countermovement jumps on bilateral force plates while wearing a multi-channel sonomyography tracking distributed wearable ultrasound system (D-WUS) on their vastus lateralis and vastus medialis oblique (Fig 1). Prior to data collection, participants provided a brief medical history and completed a clinical function assessment survey. D-WUS M-mode images were post processed using a novel composite factorization with principal component analysis method. M-mode images from the ipsilateral vastus lateralis and vastus medialis oblique were compared to characterize neuromuscular function during the countermovement jump in terms of muscle activation and muscle timing. Participants were divided into two groups based on self-reported history of knee injury within the past two years.

**Figure 1.**
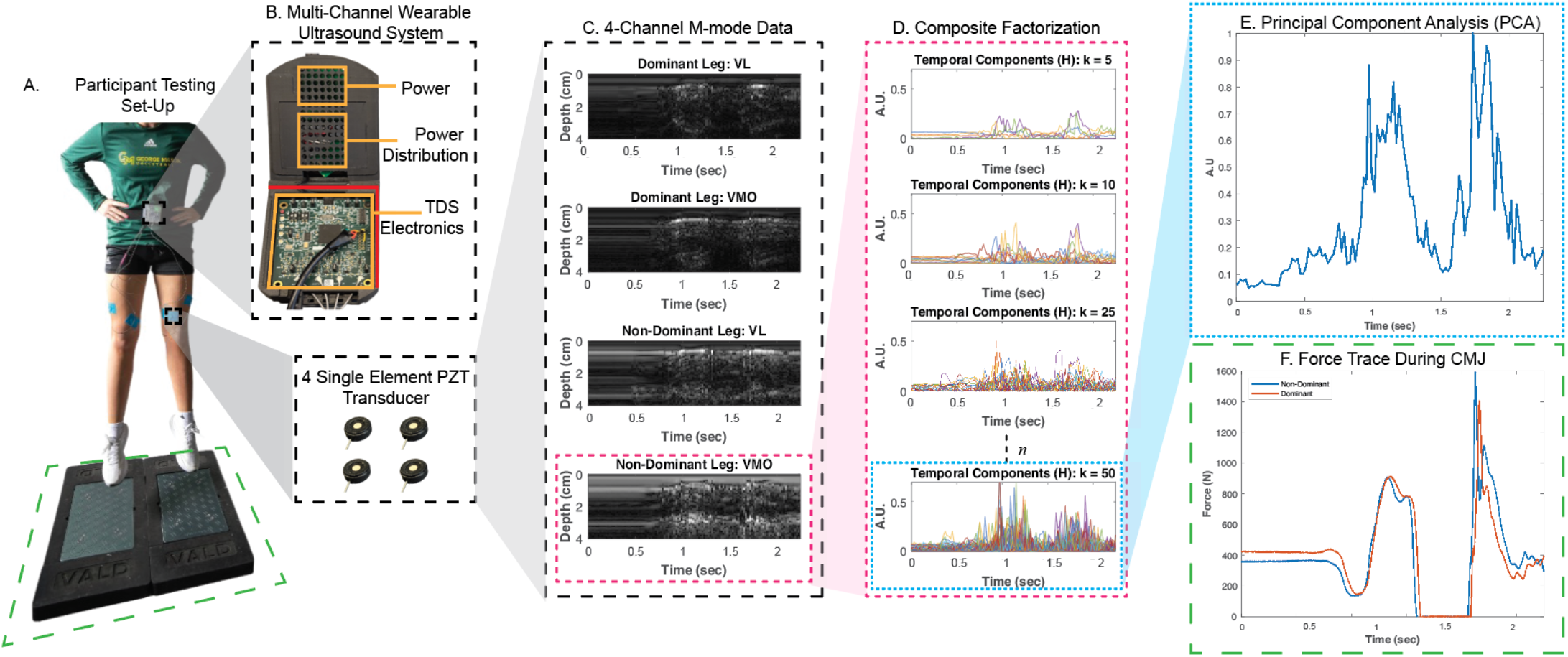
Overview of the multi-channel wearable ultrasound system and data processing workflow. (A) Participant testing setup during a countermovement jump (CMJ) on dual force plates. (B) Multi-channel wearable ultrasound system components, including power distribution, TDS electronics, and four single-element PZT transducers. (C) Representative 4-channel M-mode ultrasound data from dominant and non-dominant legs (VL and VMO muscles). (D) Composite factorization of temporal components at varying ranks (k = 5, 10, 25, 50) to extract muscle activation patterns. (E) Principal Component Analysis (PCA) of ultrasound-derived features across the CMJ cycle produces the final muscle-contraction signal, which is in agreement with (F) Ground reaction force trace during CMJ for dominant and non-dominant legs.

**Figure 2.**
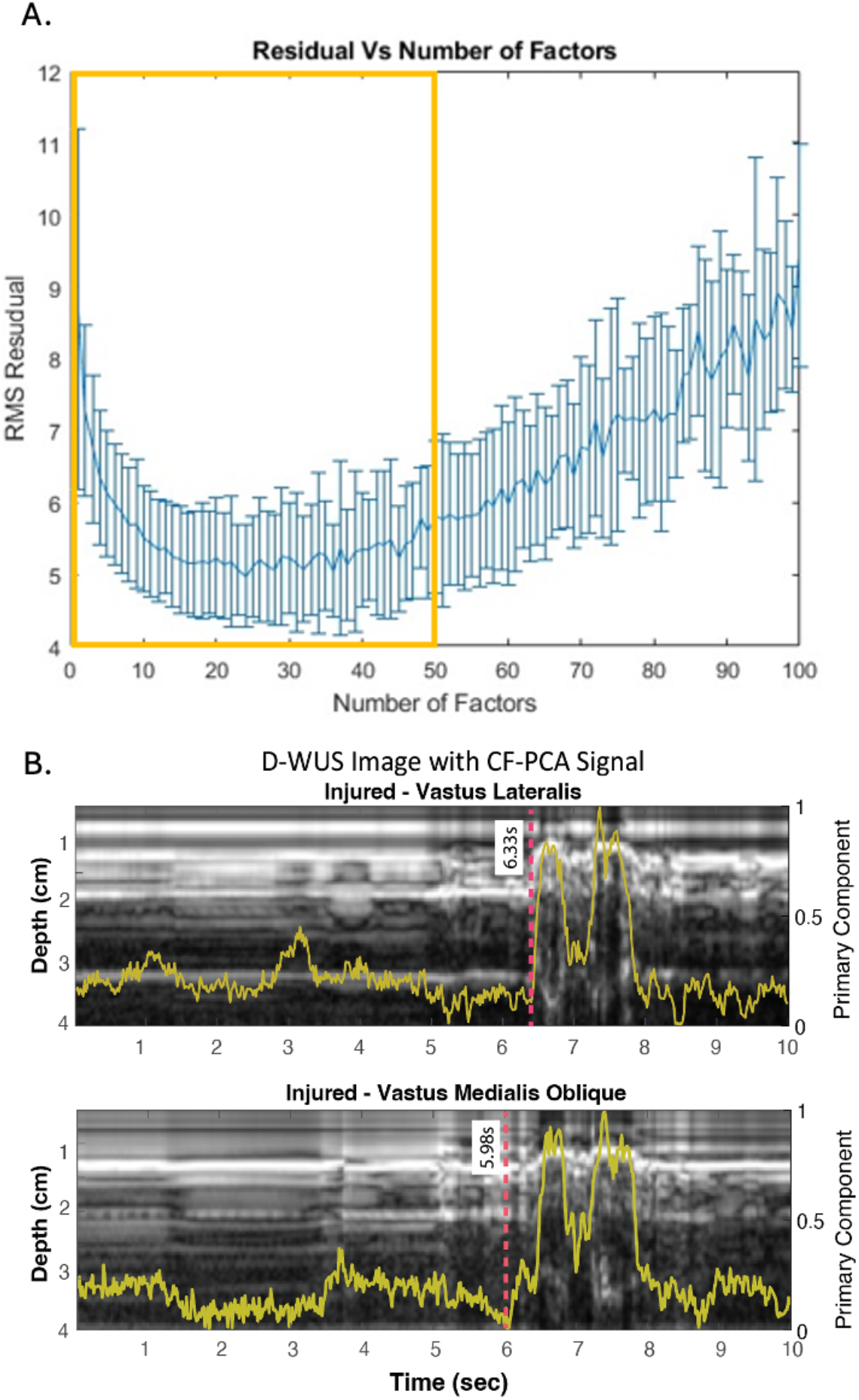
Composite factorization performance and resulting CF-PCA signal extraction. (A) RMS residual as a function of the number of NNMF factors (k). Residuals decrease initially as additional factors improve reconstruction, but beyond an intermediate range performance worsens, indicating over-factorization. Because the optimal k varies across participants and tasks, a range of factors was used for composite factorization. (B) Example multi-channel D-WUS M-mode images from the injured limb (vastus lateralis and vastus medialis oblique) with corresponding CF-PCA primary components (yellow). Dashed lines illustrate extracted timing offsets between muscle-group activation patterns.

### Participants

A convenience sample of 20 participants (mean ± SD; years: 19.8 ± 1.06, height: 176.83 cm ± 6.48 cm, weight: 77.40 kg ± 10.16 kg) were recruited for this study. All participants were current National Collegiate Athletic Association Division I women athletes, who were medically cleared to participate in collegiate athletics. Before testing began, participants in each experiment provided a brief medical history, informed consent, a short muscle function assessment survey, and indicated their dominant leg, which was determined by asking which leg they used to kick a ball [38]. They were then split into two groups based on their medical history of knee injuries, No Knee Injuries (n = 12), Knee Injuries (n= 8) (Table 1). The injured group comprised four participants with unilateral right-sided acute knee injuries (two anterior cruciate ligament (ACL) injuries with meniscal involvement, one medial patellofemoral ligament (MPFL) reconstruction, and one other right knee injury) and four with bilateral chronic knee pain (Table 2). All participants were right-leg dominant. The study adhered to the guidelines of the Declaration of Helsinki, and all procedures were approved by the University Institutional Review Board for research involving human subjects.

**TABLE 1.** Participant Demographics and injury status.

| Variable | Category | Number |
| --- | --- | --- |
| Sport | Volleyball | 11 |
|  | Basketball | 9 |
| Injury Status | Previous Knee Injury | 8 |
|  | No History of Knee Injury | 12 |
| Dominant Leg Injured | Yes | 8 |
|  | No | 0 |
| Non-Dominant Leg Injured | Yes | 4 |
|  | No | 4 |

**TABLE 2.** Injured participant pathology and injured side.

| Participant | Pathology | Injured side |
| --- | --- | --- |
| P1 | ACL + partial meniscus tear | Right |
| P2 | ACL + meniscus tear | Right |
| P3 | Right knee injury | Right |
| P4 | MPFL reconstruction | Right |
| P5 | Chronic knee pain | Bilateral |
| P6 | Chronic knee pain | Bilateral |
| P7 | Chronic knee pain | Bilateral |
| P8 | Chronic knee pain (right bursitis) | Bilateral |

All participants were right-leg dominant. ACL – anterior cruciate ligament; MPFL – medial patellofemoral ligament.

### Short Musculoskeletal Functional Assessment Survey

Prior to the start of the testing sessions, all participants were also asked to provide a brief medical history of any lower limb injuries and filled out modified lower-limb short musculoskeletal functional assessment survey (SMFA) [39]. This was used as a screening for injuries as well as an indicator of participant functional ability. SMFA ranges on a scale of 17 (healthy) to 85 (bedridden), the lower the score the better the functional ability.

### VISA-P Survey

During single session experiments, participants completed the Victorian Institute of Sport Assessment-Patella (VISA-P) scores questionnaire, which has been validated for assessing the severity of symptoms and functional ability in patients with patellar tendinopathy and PFPS [40].

### Wearable Ultrasound Sensor Placement

Each participant’s leg length was measured from their anterior superior iliac spine to the medial malleolus and the location for ultrasound was marked at 75% of their upper leg length. Clinical ultrasound images were taken using a two-dimensional B-mode ultrasound system (Clarius, L7 HD3 Linear Scanner, Vancouver, British Columbia) with a 7 cm linear transducer (frequency 4-13 MHz; axial resolution < 0.2 mm) of the participant’s vastus lateralis (VL), vastus medialis oblique (VMO). The midpoint of the clinical ultrasound transducer was marked for wearable ultrasound sensor placement. After clinical ultrasound images were acquired, the participants completed a supervised, standardized 10-minute dynamic movement warm-up. Once complete, participants were instrumented with a wearable ultrasound system and had imaging sensors placed on their dominant (D) and non-dominant (ND) VL and VMO during all experiments (Fig 1A).

### Multi-Channel Distributed Wearable Ultrasound System

A time delayed spectrometry based multi-channel wearable ultrasound (D-WUS) system was used for the collection of dynamic ultrasound data (Fig 1B) [29], [41], [42]. The D-WUS contained 4-channels using a linearly swept chirp from 3MHz to 5MHz to excite a 4.25 MHz lead zirconate titanate (PZT) transducer (SMD07T05R412WL, Steiner & Martins, Inc., Davenport, USA) in a 10-millisecond time window. Each channel consists of a single ultrasound imaging transducer which transmits the linear chirp and receives copies of the linear chirp from reflectors in the body. The transmit and receive signals are mixed and demodulated, thus allowing for ultrasound data to be sampled in the kilohertz frequency range, which significantly reduces device complexity and facilitates miniaturization. Each 10-millisecond frame of data is digitized at 40 kilo samples per second and an FFT is performed to extract reflector information to provide an A-mode signal. The spectrum magnitude identifies the strength of the reflectors, and the frequency information is then converted to depth using equation 1.

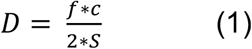

Where D is the depth of the interface, f is the frequency, c is the speed of sound, and S is the sweep rate, which is the ratio of the linear chirp range over the time window specified above.

A custom python script enables real-time signal acquisition and display of A-mode signals over time creating the M-mode image. The M-mode data is saved for further processing offline. A full description of the technology can be found in Bashatah et. al. [29].

### Countermovement Jump Testing

Participants completed countermovement jumps (CMJs) to assess lower extremity neuromuscular performance. Data were collected using dual force plates (Bertec, Bertec Corporation, Columbus OH, USA) with a sampling rate of 1000 Hz. Prior to the start of the test all participants completed a standardized warm-up. Participants performed 3 maximal effort trials of CMJs with approximately 30 seconds – 1 minute rest between each jump and were cued by the research team. CMJ variables for each leg were extracted with a custom MATLAB script (Table 3).

**TABLE 3.** Description of countermovement jump metrics.

| Variable | Abbr. | Description |
| --- | --- | --- |
| Jump Height | JH | The maximum jump height achieved, calculated using peak velocity. |
| Reactive Strength Index Modified | RSImod | Jump height divided by time to takeoff. |

*Variables were computed for each leg.*

### Composite Factorization – Principal Component Analysis

D-WUS M-modes were analyzed in post-processing using a custom MATLAB script that filtered images, applied the composite factorization principal component analysis (CF-PCA) signal processing, and computed the correlation metrics. Each m-mode frame was first normalized to its maximal pixel value and then passed through a sigmoid function. Each image was then smoothed using a Gaussian blur with a standard deviation of 2.

An unsupervised NNMF routine was implemented using the *nnmf* function in MATLAB. The *nnmf* function in MATLAB performs nonnegative matrix factorization (NNMF) on a given n × m matrix A, decomposing it into two nonnegative matrices, W (n x k) and H (k x m), where k is the desired lower rank. The product W × H provides a low-rank approximation of A, meaning that NNMF captures the dominant patterns in A while enforcing nonnegativity constraints. The factorization minimizes the root mean square residual D, given by equation 2.

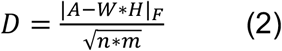

where F is the Frobenius norm. The algorithm initializes W and H randomly and updates them iteratively to minimize D. Since NNMF is not a convex problem, different runs can yield different solutions due to local minima. Sometimes, the algorithm may converge to a solution with an effective rank lower than k, which suggests the decomposition might not be optimal.

Due to NNMF randomly selecting a location for initialization and the sensitivity of choosing the number of components to extract, the number of factors was set from 1:50. The upper bound of k = 50 was selected empirically to exceed the expected number of physiologically meaningful reflector motions within the M-mode depth range, ensuring convergence without under-parameterization.

After each component was extracted, the corresponding W matrix and H matrix was concatenated to form a 1245 x m matrix of the best performing weights with the lowest root mean square residuals as determined by the NNMF algorithm.

Following NNMF, the new H matrix from the concatenation process was then reduced dimensionality using PCA to retain the first component which held the most variance. The first principal component was then selected for subsequent analyses (Fig 1E). Cross correlation and time lag was used to compare wearable ultrasound sensors within each limb for all participants: right VL vs. right VMO and left VL vs. left VMO.

The accuracy of PCA-based contraction tracking with this wearable hardware has been benchmarked against expert-annotated ground truth [57]. Among five methods (two direct signal-extraction methods, PCA alone, CF-PCA, and a supervised U-Net segmentation model), CF-PCA and PCA alone showed the closest agreement with expert fascia traces (median ρ = 0.88 for both during isometric contraction; 0.75 and 0.78, respectively, during squats; p < 0.005). PCA-based methods were recommended for tracking relative contraction patterns when labeled training data are unavailable, as in the present study.

### Statistics

Statistical analyses were conducted in MATLAB 2024b. Numerical variables were first extracted from the dataset while categorical variables were excluded. Participants were grouped by knee injury status (prior knee injury, n = 8; no history of knee injury, n = 12).

The normality of each variable was assessed using the Shapiro-Wilk test. Normally distributed variables (p ≥ 0.05) were compared between groups using Welch’s two-sample t-test (unequal variances), with effect size reported as Cohen’s d. Variables that deviated from normality (p < 0.05) were compared using the Mann-Whitney U test, with effect size reported as the rank-biserial correlation (r).

Because all participants were right-leg dominant, right- and left-limb metrics correspond to the dominant and non-dominant limbs, respectively. Right-limb, left-limb, and bilateral (right–left) VL–VMO metrics were analyzed separately.

Spearman’s rank correlations (ρ) were computed between each D-WUS coordination metric (right and left VL– VMO correlation and timing symmetry) and both clinical outcome scores (SMFA and VISA-P) across all participants (n = 20). Spearman’s correlation was selected given the small sample size and non-normal distributions identified by Shapiro-Wilk testing. This correlation analysis was exploratory: p-values were not corrected for multiple comparisons (eight tests) and should be interpreted accordingly.

All statistical tests were performed at a significance level of α=0.05. Results were compiled into a summary table, including the selected test, corresponding p-values, and normality test results for each variable.

## Results

Statistical analysis revealed a significant effect of injury status on several metrics. Right VL–VMO cross-correlation was significantly lower in participants with prior knee injury (0.816 ± 0.037) than in uninjured participants (0.862 ± 0.051; Welch’s t-test, p = 0.029, Cohen’s d = −1.02) (Fig 3C). Right VL–VMO timing symmetry was also lower in the injured group (0.728 ± 0.135 vs. 0.884 ± 0.115; p = 0.019, d = −1.26) (Fig 3D). Both represent large effects. SMFA demonstrated a significant difference between groups (29.1 ± 5.7 vs. 19.3 ± 3.6; Mann-Whitney U test, p = 0.001) (Fig 3A), indicating continued functional dysfunction associated even after significant time has passed post injury. However, VISA-P surveys did not show a significant difference between injured and non-injured participants (71.0 ± 27.0 vs. 81.3 ± 19.4; p = 0.374) (Fig 3B).

**Figure 3.**
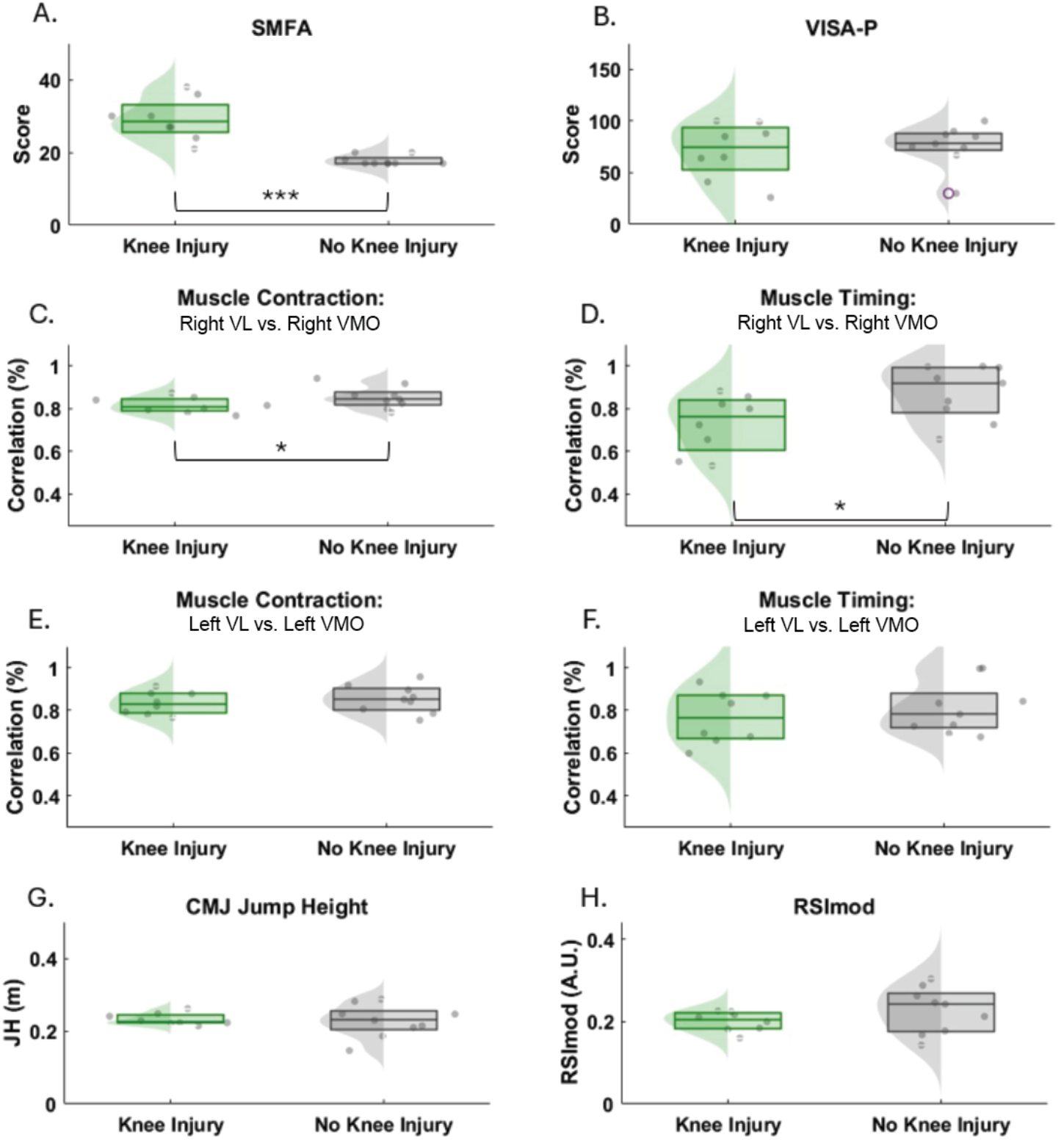
Comparison of clinical, neuromuscular, and performance measures between individuals with and without a history of knee injury. (A) SMFA scores indicate greater functional limitations in the knee injury group (**p < 0.01). (B) VISA-P scores did not differ significantly between groups. (C-D) Muscle contraction and timing correlations between vastus lateralis (VL) and vastus medialis oblique (VMO) on the right limb shows reduced coordination in the knee injury group (*p < 0.05). (E–F) Left limb muscle contraction and timing correlations did not differ significantly between groups. (G–H) Countermovement jump (CMJ) metrics, including jump height (JH), reactive strength index modified (RSImod), demonstrate no significant differences between groups. Boxplots display median, interquartile range, and individual data points with density overlays. Of the 8 injured participants, 4 sustained unilateral right-sided acute knee injuries and 4 reported bilateral chronic knee pain (Table 2).

Left VL–VMO cross-correlation did not differ significantly between groups (injured: 0.834 ± 0.053; uninjured: 0.865 ± 0.060; p = 0.247, d = −0.53), nor did left VL–VMO timing symmetry (0.767 ± 0.124 vs. 0.817 ± 0.129; p = 0.396, d = −0.40) (Fig 3E–F). Bilateral (right–left) cross-correlation (p = 0.789, d = −0.12) and timing symmetry (p = 0.817, r = 0.04) also did not differ between groups.

CMJ jump height (injured: 0.232 ± 0.016 m; uninjured: 0.226 ± 0.039 m; p = 0.612) and RSImod (0.200 ± 0.023 vs. 0.229 ± 0.048; p = 0.097) did not differ significantly between groups (Fig 3G–H).

Spearman correlations between D-WUS coordination metrics and clinical outcome scores are reported in Table 4. Right VL–VMO timing symmetry was negatively correlated with SMFA score (ρ = −0.478, p = 0.033): participants with greater self-reported musculoskeletal dysfunction showed lower between-muscle timing symmetry during the CMJ. No other D-WUS metric was significantly correlated with SMFA, and no metric was significantly correlated with VISA-P (all p > 0.05).

**TABLE 4.** Spearman correlations between D-WUS coordination metrics and clinical outcome scores (n = 20)

| D-WUS metric | vs. SMFA $\rho$ (p) | vs. VISA-P $\rho$ (p) |
| --- | --- | --- |
| Right VL–VMO correlation | –0.310 (0.184) | 0.122 (0.608) |
| Right VL–VMO timing symmetry | –0.478 (0.033)* | 0.202 (0.393) |
| Left VL–VMO correlation | 0.112 (0.638) | 0.082 (0.731) |
| Left VL–VMO timing symmetry | 0.042 (0.862) | 0.319 (0.171) |

\* $p < 0.05$ . SMFA – Short Musculoskeletal Function Assessment; VISA-P – Victorian Institute of Sport Assessment–Patella.

## Discussion

The current study addresses a gap in rehabilitation assessment by introducing a multi-site wearable ultrasound system combined with a composite signal processing pipeline to evaluate muscle-level neuromuscular coordination during dynamic movements. The primary aim of the current study was to investigate the feasibility of a time-delayed spectrometry-based wearable ultrasound system to provide objective, muscle-level assessments of neuromuscular function during dynamic tasks. By integrating this technology with countermovement jump (CMJ) testing and conventional force plate metrics, we sought to quantify global biomechanical outputs and subtle muscle activation patterns that may be overlooked with standard clinical assessments. Current findings provide preliminary evidence that prior knee injury is associated with differences in self-reported functional ability (SMFA) and right-limb VL–VMO coordination, but not in CMJ performance. Given the small cohort, these results should be regarded as a feasibility demonstration rather than evidence of clinical utility.

SMFA scores were significantly different between those with and without a previous history of knee injury (p = 0.001). The SMFA not only serves as a screening tool for detecting lower limb injuries but also provides insights into overall functional ability [43]. These self-reported measures are important because they reflect the patient’s perception of their functional limitations and pain, which are critical for tailoring individualized rehabilitation protocols [44], [45]. The difference in SMFA scores further highlights that even when objective performance metrics (such as those derived from CMJ tests) show no significant differences, subjective evaluations capture a broader spectrum of functional deficits that could affect long-term outcomes. Of note, the VISA-P survey did not differ groups in the current study. This could be due to the VISA-P survey’s questions being focused how the athlete’s injury is affecting playing time and not functional ability [40]. The athletes in the current study were fully cleared to participant in all aspects of their sport.

The CMJ test is widely used in sports science to monitor neuromuscular performance, fatigue, and readiness to return to play [46], [47]. In the current study, CMJ performance metrics (JH and RSImod) were not different between groups. Paterno et al., reported that female athletes who underwent ACL reconstruction demonstrated lower limb asymmetries during jumping tasks after their return to sport (+2 years) [48]. However, their measures used focused solely on vertical ground reaction force during a drop vertical jump on bilateral force plates [48]. Further, there were biomechanical differences between CMJ and vertical drop jump landing mechanics [49]. CMJ landing mechanics were not analyzed as part of this study.

While the CMJ metrics showed no difference between the injured and non-injured group the D-WUS results showed muscle level differences. The current analysis indicated that both the correlation and timing of activation between these muscle groups differed significantly between injured and non-injured athletes’ right VL and VMO. This finding is particularly relevant because proper coordination between the VL and VMO is critical for patellar tracking and knee stability [16], [50], [51]. In athletes with knee injuries, the altered timing and reduced correlation may reflect compensatory neuromuscular adaptations, potentially contributing to chronic conditions such as patellofemoral pain syndrome [50].

Left-limb VL–VMO coordination did not differ significantly between groups. This is consistent with the injury distribution of this cohort: four of the eight injured participants had unilateral right-sided acute injuries, and compensatory adaptations would be expected primarily in the injured limb. The lateralized pattern of significant findings should therefore be interpreted in light of the injury distribution rather than as a broad physiological effect. Future studies should include cohorts with left-sided and bilateral injuries to characterize contralateral compensation.

Traditional rehabilitation protocols rely on subjective assessments, visual observation, and force plate analysis to evaluate overall neuromuscular performance [47], [52]. The current findings suggest that self-reported functional assessments (SMFA), which measures difficulty in performing activities of daily living, are consistent with muscle-level coordination metrics obtained from the wearable ultrasound system. However, functional assessments that are based on performance and quantitative performance assessments show a different result. Although force plates provide objective measures of global biomechanics, they do not capture detailed, muscle-level activation patterns. Surface electromyography (sEMG) has been employed to monitor muscle activity; however, its limitations, such as signal noise, muscle crosstalk, and inability to assess deep-seated muscles, restrict its effectiveness during dynamic tasks [18], [19], [53].

Right VL–VMO timing symmetry was negatively correlated with SMFA score (ρ = −0.478, p = 0.033), providing preliminary evidence linking D-WUS coordination metrics to self-reported functional status. No significant correlations were found with VISA-P, possibly because that instrument targets patellar tendon symptoms specifically. These correlations should be interpreted cautiously given the sample size and the number of comparisons, and replication in larger cohorts is needed.

In contrast, multi-channel D-WUS enables direct, real-time imaging of muscle activity at multiple sites simultaneously. Through the CF-PCA signal processing technique information that is directly related to the muscle movement can be seen and extracted. Iverson et al. used a similar NNMF algorithm to extract information from clinical ultrasound images while participants were performing a walking task [34]. Results showed that signals extracted from the clinical 2D images correlated to phase of the gait cycle, which could be used in conjunction with machine learning to control wearable devices [34]. However, due to the need for generalizable data in a timely fashion CF-PCA is well suited to this application because without using machine learning it decomposes the complex ultrasound data into dominant activation patterns, making the process computationally efficient and suitable for near real-time analysis. Additionally, due to the multi-factorization done in the CF-PCA, the variability in the image is retained minimizing data loss that is possible with traditional NNMF methods [54].

The coordination metrics reported here depend on the relative shape and timing of contraction signals between muscles, not on absolute fascia depth. A separate benchmarking study using the same wearable hardware found that CF-PCA and PCA alone tracked relative contraction patterns with comparable fidelity to expert annotation (median ρ = 0.88 for both during isometric contraction; 0.75 and 0.78 during squats) [57]. This suggests that the between-group differences observed here reflect information in the D-WUS signal rather than a property specific to the CF-PCA formulation. Supervised segmentation (U-Net) achieved lower absolute depth error in that study and may be preferable once labeled training data are available for dynamic tasks such as the CMJ.

If confirmed in larger cohorts, these findings may have implications for rehabilitation. Ensuring that neuromuscular control is fully restored during the late stages of rehabilitation is paramount to reducing the risk of re-injury and facilitating a safe return to sport [5], [55]. Traditional strength assessments may not detect lingering neuromuscular deficits, leading to premature return-to-play decisions [56]. By enabling direct measurement of muscle-level activation and coordination, D-WUS may offer a useful tool for clinicians to evaluate recovery. For instance, the significant differences in dominant VL-VMO correlation and timing observed suggest that, if confirmed in larger cohorts, monitoring these parameters could serve as an early indicator of persistent deficits. Integrating D-WUS with conventional load monitoring strategies, such as those based on CMJ metrics, could provide a more holistic framework for evaluating rehabilitation progress.

Moreover, the ability to monitor these parameters over multiple sessions could offer valuable insights into the longitudinal effects of rehabilitation protocols. By tracking changes in muscle-level function over days or weeks, clinicians could adjust interventions more precisely, thereby optimizing recovery and performance.

This study has several limitations. The sample size was relatively small and restricted to NCAA Division I women athletes who were cleared for activity, which may limit the generalizability of the findings. Additionally, the heterogeneity of injury types, ranging from acute injuries to chronic knee pain and other knee-related conditions, introduces variability that could confound the interpretation of neuromuscular metrics. The injured group comprised both unilateral right-sided acute injuries and bilateral chronic knee pain (Table 2); the predominance of right-sided acute injuries likely explains the lateralized pattern of significant findings, and the small subgroups precluded analysis by pathology type. With eight injured participants, the study was also underpowered to detect moderate effects, so non-significant results should not be interpreted as evidence of no difference. Future research should aim to include larger, more diverse cohorts, as well as incorporate male athletes and athletes from various sports to understand how sport-specific mechanics influence neuromuscular function at the muscle level.

The present study was intended as an initial investigation of wearable ultrasound–derived muscle coordination metrics during dynamic tasks. While statistically significant differences were observed in muscle-level coordination measures, interpretation of non-significant differences in global performance metrics should be made cautiously. Future studies with larger cohorts will be valuable for further characterizing relationships between wearable ultrasound features and conventional biomechanical measures across broader athlete populations.

Another important area for future investigation is the refinement of the signal processing pipeline. While CF-PCA provides a means for analyzing the ultrasound data, further improvements in these methods could enhance the robustness, sensitivity, and specificity of muscle-level assessments. This study used a single-session protocol. Before D-WUS can support clinical decisions, future work must establish sensor placement repeatability, coupling stability during dynamic movement, robustness to motion artifact, and inter-session reliability of the coordination metrics, followed by longitudinal studies across rehabilitation.

## Conclusions

In conclusion, the current study provides preliminary evidence that wearable ultrasound technology, combined with advanced signal processing, can capture muscle-level neuromuscular function during dynamic tasks. Multi-channel D-WUS detected right-limb differences in VL–VMO coordination between injured and non-injured athletes that were not reflected in conventional force plate metrics. These findings suggest that wearable ultrasound may, with further validation, provide clinicians an objective, real-time means of assessing neuromuscular recovery during late-stage rehabilitation. Future work with larger, more diverse cohorts will be needed to establish its reliability and clinical utility.

## Data Availability

All data produced in the present study are available upon reasonable request to the authors.

## Acknowledgment

The authors would like to acknowledge that these efforts were sponsored by the Government under Other Transactions Number W81XWH-15-9-0001.

